# Regional variations in Antibiotic Consumption in Tanzania: A Geospatial Analysis of eLMIS Data for Targeted Stewardship

**DOI:** 10.1101/2025.10.22.25338546

**Authors:** William Reuben, Florah Boniface Makenya, Raphael Z. Sangeda, Daudi Ignasy Msasi, Selesitine Ngoma

**Affiliations:** Department of Health, Social Development and Nutrition, President’s Office, Regional Administration and Local Government, Tanzania; School of Nursing and Public Health, University of Dodoma, Dodoma, Tanzania; Department of Pharmaceutical Microbiology, Muhimbili University of Health and Allied Sciences, Tanzania; Pharmaceutical Services Unit, Ministry of Health, Dodoma, Tanzania; Department of Public Health and Community Nursing, University of Dodoma, Tanzania

**Keywords:** antibiotic consumption, antimicrobial stewardship, geospatial analysis, eLMIS, Tanzania, AWaRe

## Abstract

**Background:** Understanding regional variations in antibiotic consumption is critical for guiding antimicrobial stewardship (AMS) in low- and middle-income countries. Although national antibiotic use data exist for Tanzania, spatial disparities across regions remain poorly characterized.

**Methods:** This retrospective analysis utilized facility-level antibiotic supply data from the electronic Logistics Management Information System (eLMIS), using data from July 2020 to June 2024. Antibiotic consumption was expressed as defined daily doses per 1,000 inhabitants per day (DID) following the WHO ATC and AWaRe classifications. Regional- and facility-level variations were analyzed and visualized using descriptive and geospatial mapping approaches. Temporal trends and national forecasts were modeled using ARIMA and polynomial regression.

**Results:** A total of 1.63 million facility-level records were analyzed. National antibiotic consumption increased from 142.4 to 182.8 DID between 2020–2021 and 2023–2024 (a 28% rise). Dispensaries accounted for 71% of the total DID, highlighting the dominant role of primary care in antibiotic distribution. Dar es Salaam (37.9 DID), Ruvuma (34.5 DID), and Lindi (34.0 DID) showed the highest cumulative consumption, whereas Katavi (13.7 DID) and Geita (15.4 DID) recorded the lowest. The access category antibiotics comprised ≥ 60% of the total consumption, Watch category antibiotics comprised 35–40%, and Reserve category antibiotics comprised ≤ 0.1%. The forecasting models project continued growth, reaching approximately 215 DID by 2027.

**Conclusions:** Antibiotic consumption in Tanzania exhibits a rising trend characterized by pronounced regional disparities, particularly in the coastal and southern regions. The analysis emphasizes the importance of tailoring AMS programs to local contexts and illustrates how digital supply chain platforms can inform geographically targeted interventions in resource-constrained settings. Strengthening AMS at the primary care level and prioritizing high-consumption regions are essential steps.

## Introduction

Antimicrobial resistance (AMR) is a growing global health threat, primarily driven by the misuse and overuse of antibiotics [1,2]. Understanding antibiotic consumption patterns is critical for guiding stewardship (AMS) efforts and ensuring the rational use of vital medicines [1]. Traditionally, antibiotic utilization has been monitored at the national or regional level and expressed as defined daily doses per 1,000 inhabitants per day (DID). Such metrics provide valuable insights into overall trends and have been widely used in national AMR surveillance programs, including those in Tanzania [3].

However, national-level estimates often mask substantial regional differences in antibiotic consumption, which may contribute to the emergence of localized AMR [2]. A landmark global study by Browne et al. [4] employed spatial modeling to reveal striking disparities in antibiotic use across 204 countries, underscoring the importance of geospatial approaches in guiding antibiotic stewardship. Similarly, neighborhood-level analyses in high-income settings have shown that geospatial mapping can pinpoint hotspots of antibiotic misuse and resistance [5,6]. For example, in Italy, variations in governance and healthcare quality have been linked to regional differences in antibiotic use, illustrating the complex interplay between social and health system factors that shape these patterns.

Despite these advances, a critical gap remains in the geospatial analysis of antibiotic use in low- and middle-income countries (LMICs). In Tanzania, although national antibiotic consumption estimates are available [3,7–9], no study to date has mapped regional differences in antibiotic consumption, despite Tanzania having well-documented variations in healthcare infrastructure and disease burden [10]. Urban centers, such as Dar es Salaam, may have higher access to antibiotics and distinct prescribing practices compared to rural regions. These disparities necessitate locally tailored AMS interventions aligned with the One Health Framework [11].

Despite multiple national consumption analyses, none have spatially quantified variations that can guide decentralized AMS and procurement. Tanzania’s Electronic Logistics Management Information System (eLMIS) offers a unique opportunity to address this gap. By leveraging this dataset, our study aimed to quantify and map regional variations in antibiotic consumption across Tanzania’s regions, identify potential hotspots of overuse or underuse, and provide data-driven insights that can inform region-specific AMS interventions and resource allocation. Our ultimate objective was to generate actionable evidence that supports Tanzania’s national AMR Action Plan and guides equitable targeted AMS initiatives across diverse regional contexts.

## Methodology

### Study Design and Setting

This retrospective, facility-level study analyzed antibiotic consumption patterns across Tanzania’s 26 administrative regions. Data were obtained from the eLMIS, which records routine reports on the distribution of medicines from public and private healthcare facilities nationwide.

### Data Source and Timeframe

Monthly antibiotic distribution data were extracted from eLMIS for the period from July 2020 to June 2024, covering the fiscal years 2020/2021 – 2023/2024. The dataset included the quantities of antibiotics supplied to each facility, classified by product name, formulation, and strength.

### Antibiotic Classification and Measurement

Antibiotics were categorized according to the World Health Organization Anatomical Therapeutic Chemical (ATC) classification system. Consumption was standardized using defined daily doses (DDD) and expressed as DDD per 1,000 inhabitants per day (DID), calculated as follows:

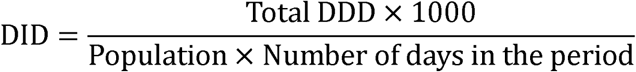

Where *Total DDD* represents the aggregated number of DDDs distributed, and *population* refers to the estimated regional population based on official census projections. Antibiotics were further grouped by ATC Level 4 class and WHO AWaRe category (Access, Watch, Reserve) to assess prescribing balance. Regional population estimates were derived from the 2022 Tanzania Census projections [12].

### Data Analysis

The analyses combined descriptive statistics, geospatial mapping, and time-series modeling. Descriptive summaries captured the national and regional trends in antibiotic consumption and facility-level distribution volumes.

All analyses were conducted in R version 4.3.2, using the packages sf, ggplot2, and forecast. These packages were used to link regional DID data with national administrative boundaries (ADM1 shapefiles from the Humanitarian Data Exchange) and to generate thematic maps of antibiotic distribution intensity.

Time-series analyses assessed annual trends and projected national consumption for 2025–2027 using both Autoregressive Integrated Moving Average (ARIMA [1,1,0]) and second-degree polynomial models. Model fit was evaluated by visual inspection and the Akaike Information Criterion (AIC). The polynomial model (R² = 0.97) showed a slightly better predictive accuracy, capturing the post-2022 acceleration in consumption.

### Ethical Considerations

This study utilized secondary, routinely collected data from the eLMIS, which the Tanzanian Ministry of Health maintains for supply chain and public health monitoring. No patient-level data were available. Ethical approval was obtained from the University of Dodoma Institutional Review Board (Ref: MA.84/261/87/172). Permission to use eLMIS data was obtained from the Ministry of Health and the President’s Office – Regional Administration and Local Government (PORALG). All data were anonymized, aggregated, and handled in accordance with national data-protection regulations.

## Results

A total of 1,633,110 facility-level antibiotic distribution records were retrieved from eLMIS for the period from July 2020 to June 2024. These data represent aggregated supplies to healthcare facilities rather than individual patient prescriptions.

Total antibiotic consumption, expressed as defined daily doses per 1,000 inhabitants per day (DID), showed a consistent upward trend over the four years (Table 1). The summed national DID increased from 142.37 in 2020–2021 to 182.79 in 2023–2024, reflecting a 28.3% rise in antibiotic supply. Facility reporting also improved over time, with the number of records increasing from 315,707 to 494,173.

**Table 1:** Annual antibiotic consumption and number of facility-level records, 2020–2024.

| **Fiscal Year** | **Sum of DID** | **No. of Facility Records** | **Mean DID (± SD)** |
| --- | --- | --- | --- |
| 2020–2021 | 142.37 | 315,707 | 4.60 ± 2.32 |
| 2021–2022 | 163.76 | 393,533 | 5.29 ± 2.73 |
| 2022–2023 | 162.01 | 429,697 | 5.23 ± 2.68 |
| 2023–2024 | 182.79 | 494,173 | 5.90 ± 2.97 |
| **Total / Overall** | **650.92** | **1,633,110** | **5.26 ± 2.68** |

### Antibiotic Distribution by Facility Level

Analysis of antibiotic distribution by customer level revealed that dispensaries received the largest share of antibiotics, accounting for a cumulative 459.71 DID (70.6%) during the study period (Table 2). This was followed by district hospitals (56.23 DID; 8.6%) and health centers (54.44 DID; 8.4%). Collectively, these three primary-level facility types accounted for over 87% of all antibiotic distributions recorded in the eLMIS dataset, underscoring the central role of primary healthcare facilities in national antibiotic use.

**Table 2:**
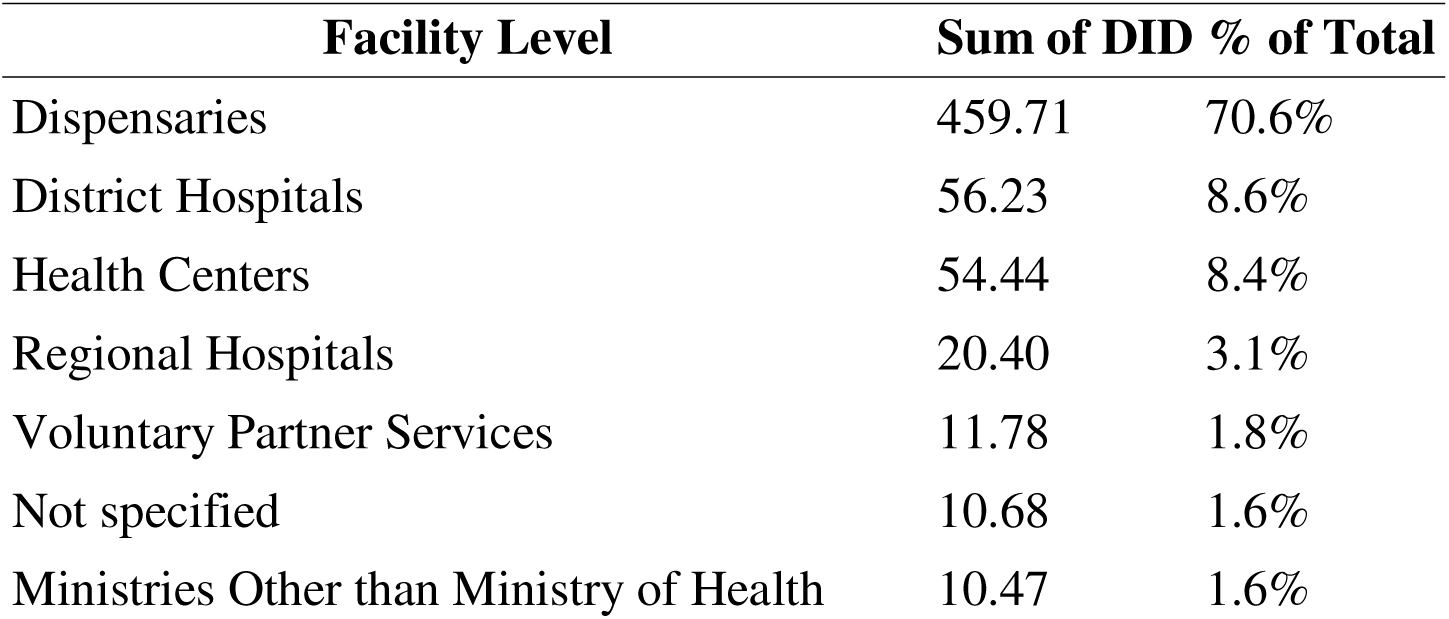
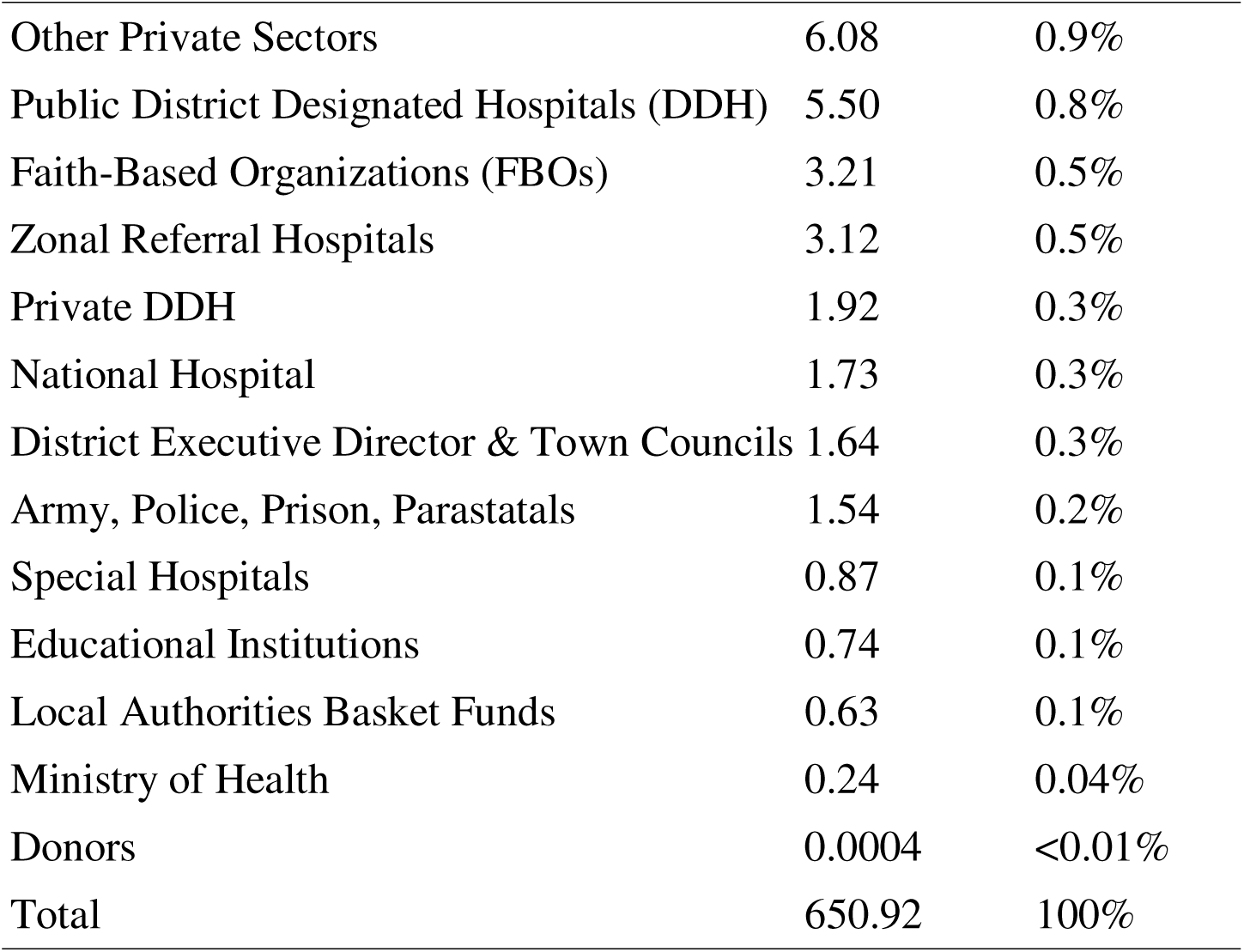
Cumulative Sum of Antibiotic Distribution (DID) by Customer Level (Facility Type), 2020–2024.

Other facility types, including regional hospitals (20.40 DID; 3.1%), voluntary partner (VP) services (11.78 DID; 1.8%), and zonal referral hospitals (3.12 DID; 0.5%), contributed smaller but notable shares. Specialized and institutional facilities, such as faith-based organizations (3.21 DID; 0.5%), national hospitals (1.73 DID; 0.3%), and army, police, prisons, and parastatal services (1.54 DID; 0.2%), accounted for a limited proportion of the total distribution.

The lowest volumes were recorded for educational institutions (0.74 DID), local authorities (0.63 DID), and donor-funded facilities (0.0004 DID).

### Regional Antibiotic Consumption Patterns

Antibiotic consumption patterns exhibited marked regional variation across Tanzania (Figure 1). Dar es Salaam recorded the highest cumulative antibiotic consumption at 37.9 DID, followed by Ruvuma (34.5 DID) and Lindi (34.0 DID). Other regions with elevated use included Iringa (30.6 DID), Njombe (29.7 DID), and Mtwara (29.2 DID). In contrast, the lowest cumulative values were observed in Katavi (13.7 DID), Geita (15.4 DID), and Simiyu (15.7 DID) across the four-year study period.

**Figure 1:**
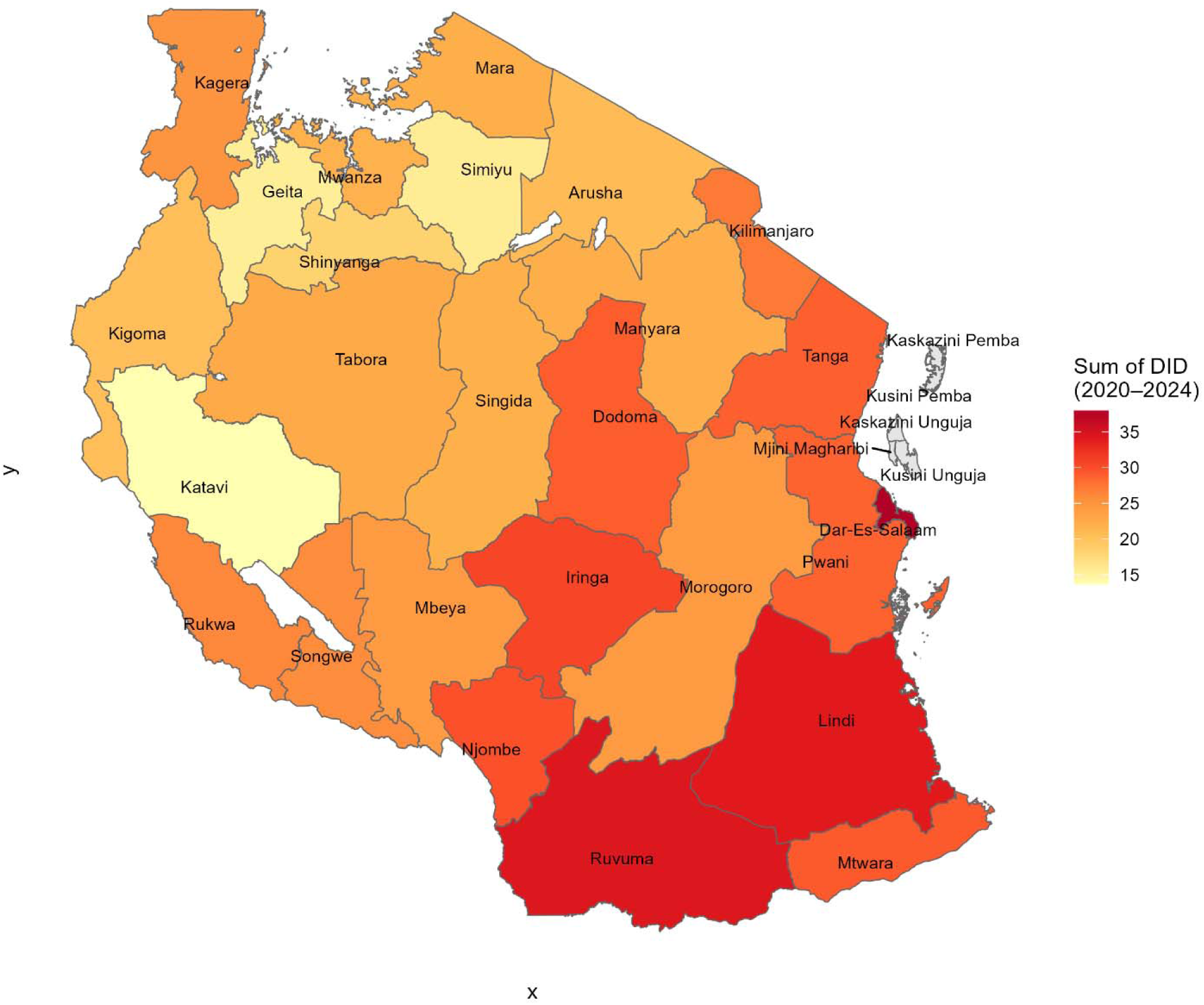
Regional antibiotic distribution in Tanzania (2020–2024). Cumulative antibiotic distribution across 26 regions, expressed as total defined daily doses per 1,000 inhabitants per day (DID), based on facility-level supply data from the eLMIS. Geographic boundaries were derived from Administrative Level 1 (ADM1) shapefiles provided by the National Bureau of Statistics (NBS) Tanzania website https://www.nbs.go.tz/statistics/topic/gis, which is openly available under the CC-BY 4.0 license.

Although Dodoma had the highest number of facility-level records (99,477), it did not rank among the top consumers. Conversely, regions such as Dar es Salaam, Ruvuma, and Lindi have relatively high consumption levels, with smaller reporting volumes.

As summarized in Supplementary Table 1, cumulative DID values varied widely, ranging from 37.9 DID in Dar es Salaam to 13.7 DID in Katavi, highlighting pronounced geographical heterogeneity in antibiotic use.

### Antibiotic Consumption by Molecule

During the study period, antibiotic distribution in Tanzania was dominated by a limited number of agents (Figure 2). The top 10 antibiotics accounted for more than 90% of the total defined daily doses (DDD), with amoxicillin, sulfamethoxazole + trimethoprim, doxycycline, ciprofloxacin, and erythromycin together contributing nearly three-quarters of all distributed doses. Most of these agents belong to the Access group.

**Figure 2:**
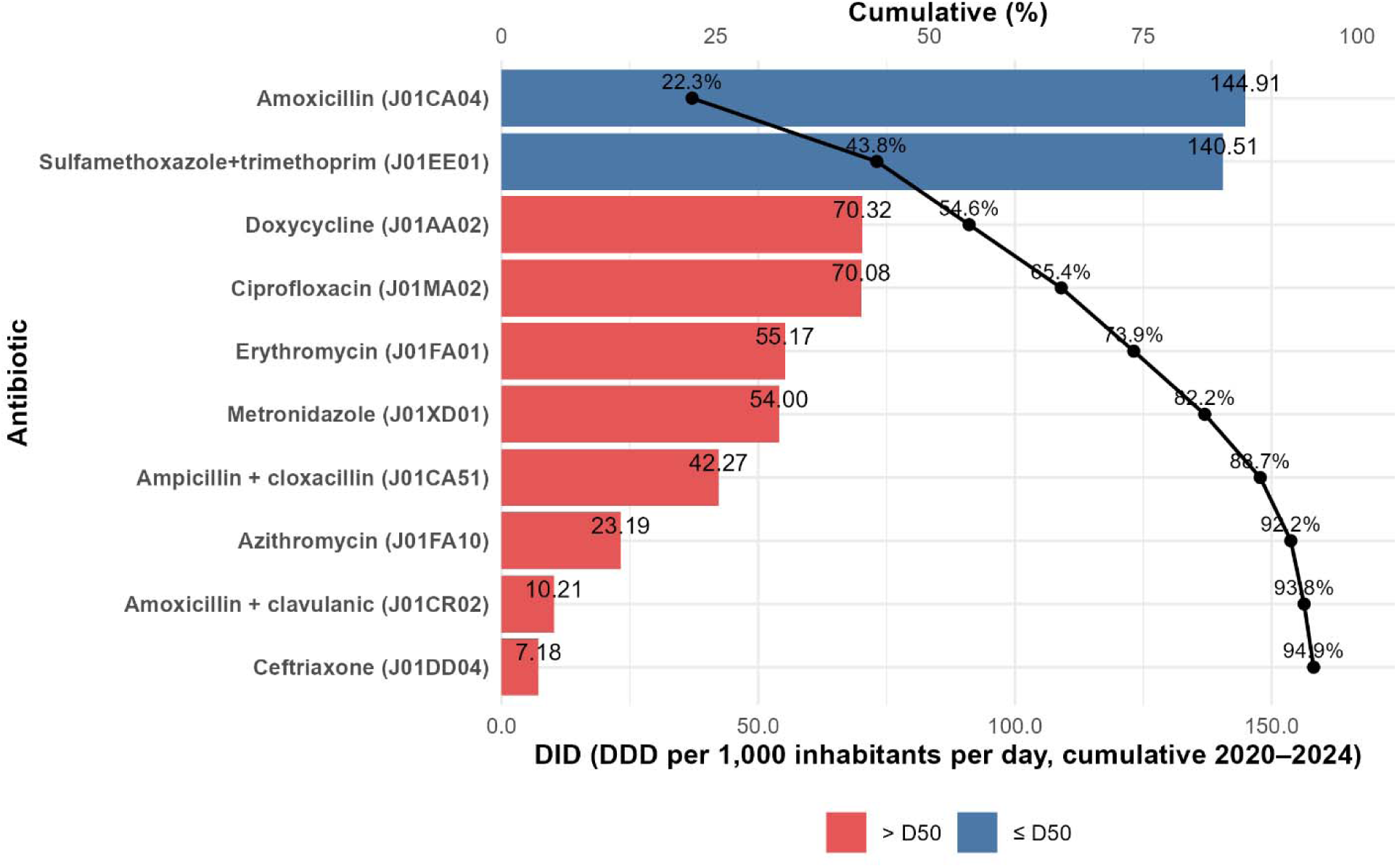
Top 10 antibiotics by defined daily doses (DDD per 1,000 inhabitants per day), Tanzania, 2020–2024. Bars represent cumulative antibiotic distribution (DID) by molecule; the overlaid line and points indicate the cumulative percentage contribution. Bar colors denote D50 membership (≤50% cumulative for each rank). Source: eLMIS facility-level supply data. The complete list is provided in Supplementary Table 2.

The detailed distribution of all antibiotics, with a minimum of 60 entries and their cumulative contributions, is presented in Supplementary Table 2.

### AWaRe distribution by year

The distribution of antibiotic consumption by the WHO AWaRe category showed a consistent predominance of the Access category antibiotics throughout the study period (2020–2024) (Figure 3). Access agents accounted for 55.4–63.0% of total DID, reaching 61.0% in 2023–2024. Antibiotics accounted for 37.0–44.5% of annual consumption, with a peak of 39.0% recorded in the 2023–2024 period. Reserve category of antibiotics contributed minimally (≤0.1% each year, with a rate of 0.04% from 2023 to 2024).

**Figure 3:**
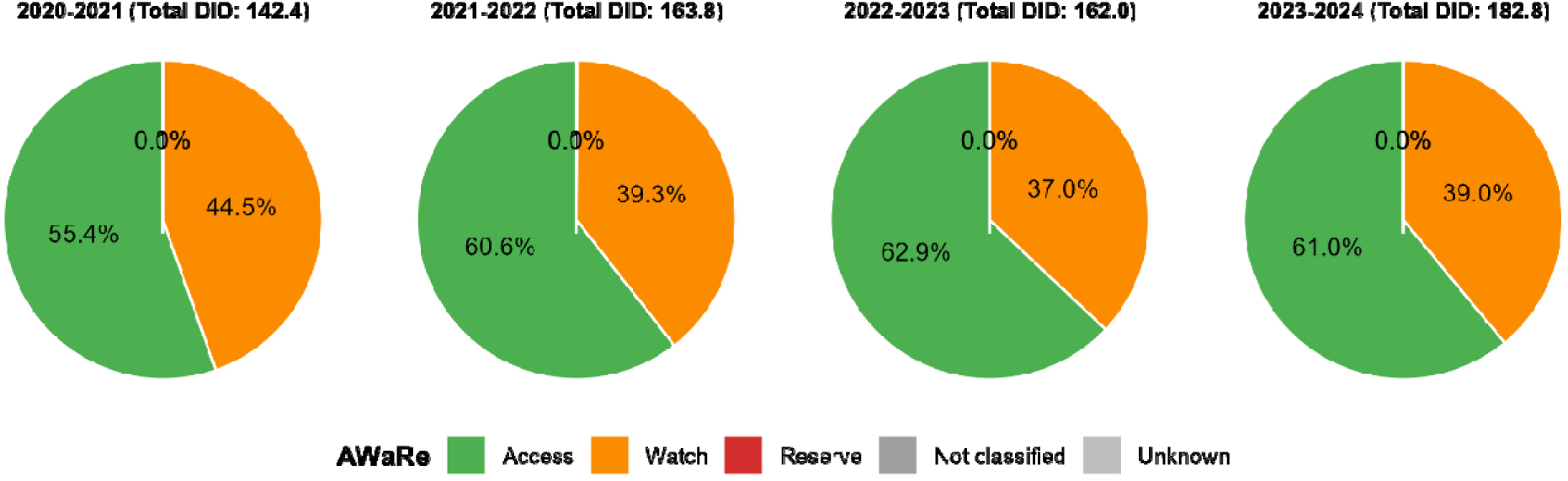
AWaRe distribution by year, Tanzania, 2020–2024. Each pie chart depicts the annual share of antibiotic consumption (DID) in the AWaRe category. Colors: Access (green), Watch (orange), Reserve (red), Not classified (gray), Unknown (light gray). Source: eLMIS facility-level supply data.

The proportion of Access agents surpassed the WHO-recommended benchmark of≥60% from 2021 to 2022.

### Dosage forms

Antibiotic distribution across 2020–2024 was dominated by tablets, which accounted for the largest share of the total DID throughout the study period (Figure 4). However, their relative contribution declined from 60.1% in 2020–2021 to 50.4% in 2023–2024. In contrast, capsules increased from 29.1% to 34.6%, syrups from 8.3% to 10.9%, and injections from 2.5% to 4.1% over the same period.

**Figure 4:**
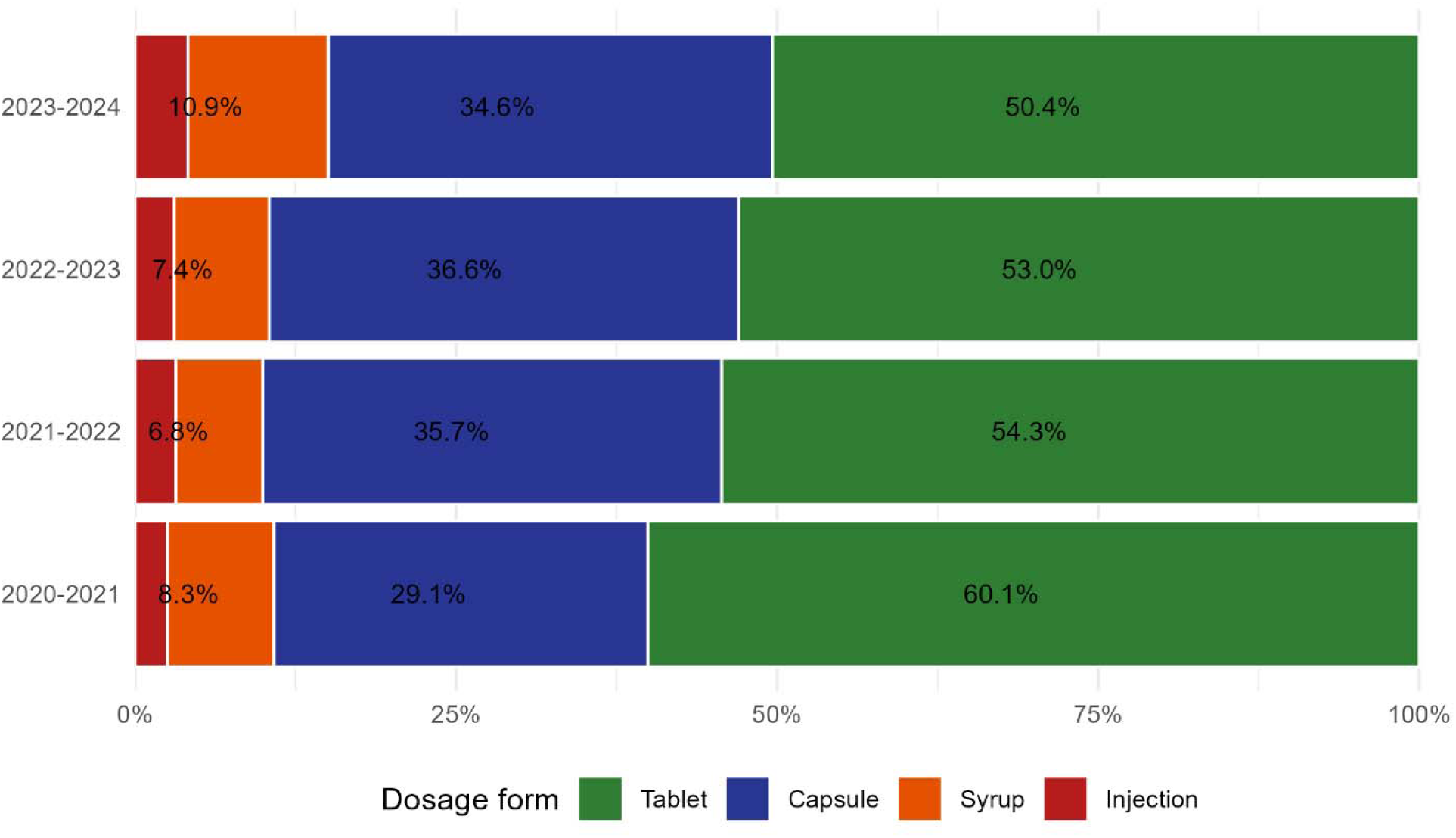
Dosage-form distribution of annual antibiotic consumption, Tanzania, 2020– 2024. Stacked bars show the proportion (%) of the total defined daily doses per 1,000 inhabitants per day (DID) by dosage form for each study year.

Overall, antibiotic supply rose from 142.37 to 182.79 DID.

### ATC Level 3 distribution

Cumulative antibiotic distribution from 2020 to 2024 was dominated by penicillins (J01C), which accounted for 32.6% of the national total (Figure 5). This was followed by sulfonamide– trimethoprim combinations (J01E; 21.6%), macrolides, lincosamides, streptogramins (J01F; 12.2%), quinolones (J01M; 10.9%), tetracyclines (J01A; 10.8%), and other antibacterials (J01X; 9.1%). Together, these six major classes represented 97.1% of all antibiotics distributed during the study period, with the top five classes alone contributing 88.1% of the total DID.

**Figure 5:**
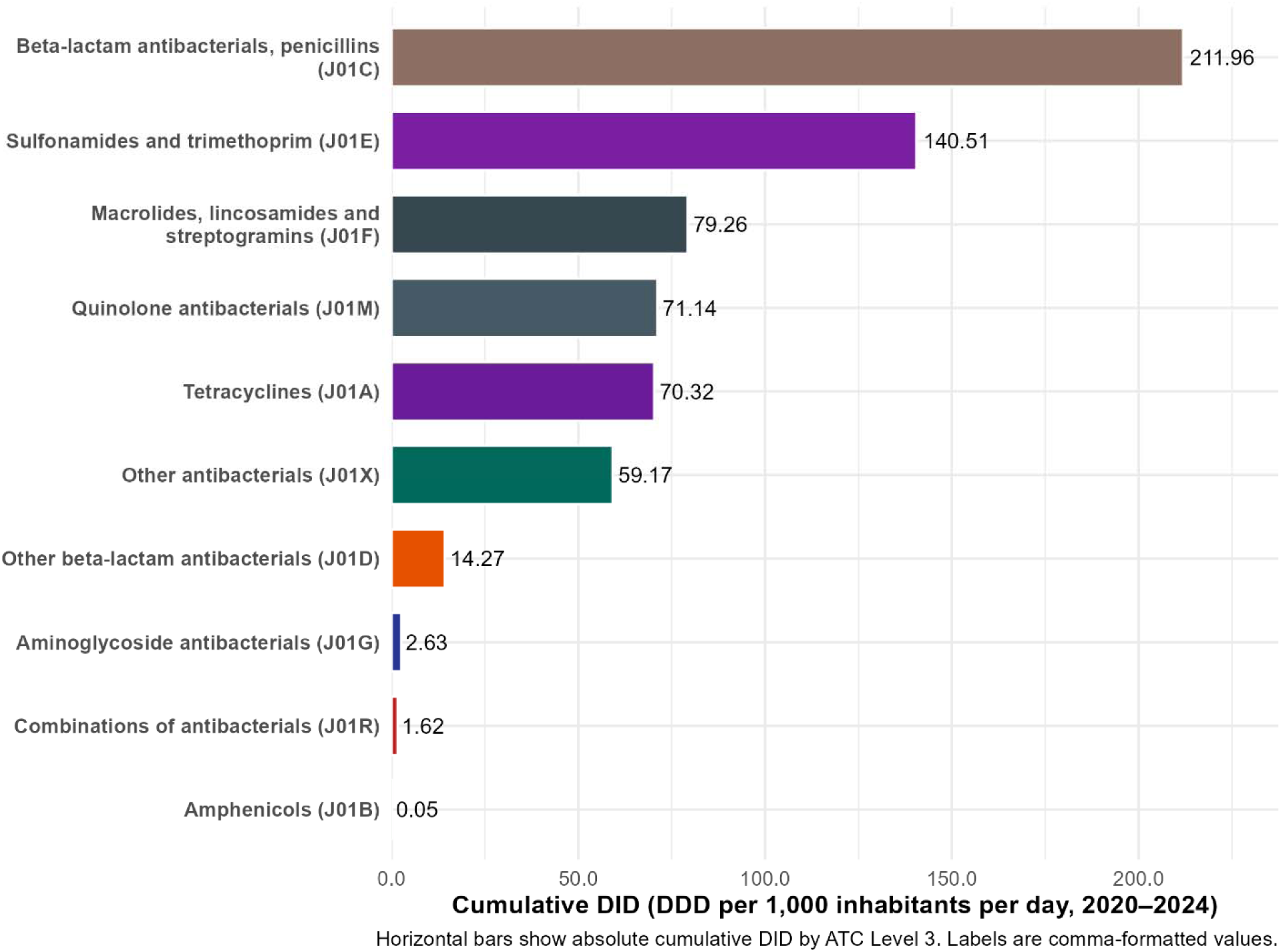
ATC Level 3 distribution of cumulative antibiotic consumption, Tanzania, 2020– 2024. Horizontal bars display the total defined daily doses per 1,000 inhabitants per day (DID) by therapeutic class, with labels showing absolute values. Source: eLMIS facility-level supply data.

The minor classes included other β-lactams (J01D; 2.2%), aminoglycosides (J01G; 0.4%), combinations (J01R; 0.2%), and amphenicols (J01B; <0.1%). This distribution highlights Tanzania’s strong reliance on a limited set of broad-spectrum antibiotic classes, consistent with national treatment guidelines and the predominance of Access agents observed in the AWaRe analysis.

### ATC Level 4 distribution

At the ATC Level 4 classification, penicillins with extended spectrum (J01CA) were the leading antibiotic class in all study years, increasing from 48.48 DID in 2020–2021 to 50.75 DID in 2023–2024 (class total 188.54 DID; ∼29% of all antibiotics distributed) (Supplementary Table 3). This was followed by sulfonamide–trimethoprim combinations (J01EE; 140.51 DID; ∼22%), which declined from 44.91 to 30.70 DID over the same period.

Other major contributors included macrolides (J01FA; 79.26 DID; ∼12%), which more than doubled from 11.99 to 26.93 DID, fluoroquinolones (J01MA; 71.14 DID; ∼11%), and tetracyclines (J01AA; 70.32 DID; ∼11%), which peaked in 2022–2023 before a modest decline. Imidazoles (J01XD) rose steadily from 6.89 to 21.61 DID (class total 56.39 DID; ∼9%).

Smaller but notable growth was observed in β-lactam/β-lactamase inhibitor combinations (J01CR; class total 10.21 DID) and third-generation cephalosporins (J01DD; 8.85 DID), while Reserve-leaning parenteral classes such as carbapenems (J01DH; 0.04 DID) and glycopeptides (J01XA; 0.009 DID) remained negligible.

Overall annual totals increased from 142.37 DID in 2020–2021 to 182.79 DID in 2023–2024, underscoring both the rising volume and diversity of antibiotic consumption during the study period.

### Forecasting of National Antibiotic Consumption (2025–2027)

Time-series modeling using an ARIMA (1,1,0) model and second-degree polynomial fits was applied to project antibiotic consumption for the years 2025–2027, based on the trend observed from 2020 to 2024. Both models predicted a continued national increase in antibiotic use, with the polynomial model demonstrating a slightly superior fit (R² = 0.97) and a better representation of the post-2022 acceleration pattern.

Forecasts indicate that national antibiotic consumption will rise from 182.79 DID in 2023–2024 to approximately 195.6 DID in 2024–2025, 205.8 DID in 2025–2026, and 215.2 DID in 2026– 2027, assuming no major AMS interventions or supply disruptions occur (Supplementary Table 4).

**Figure 6:**
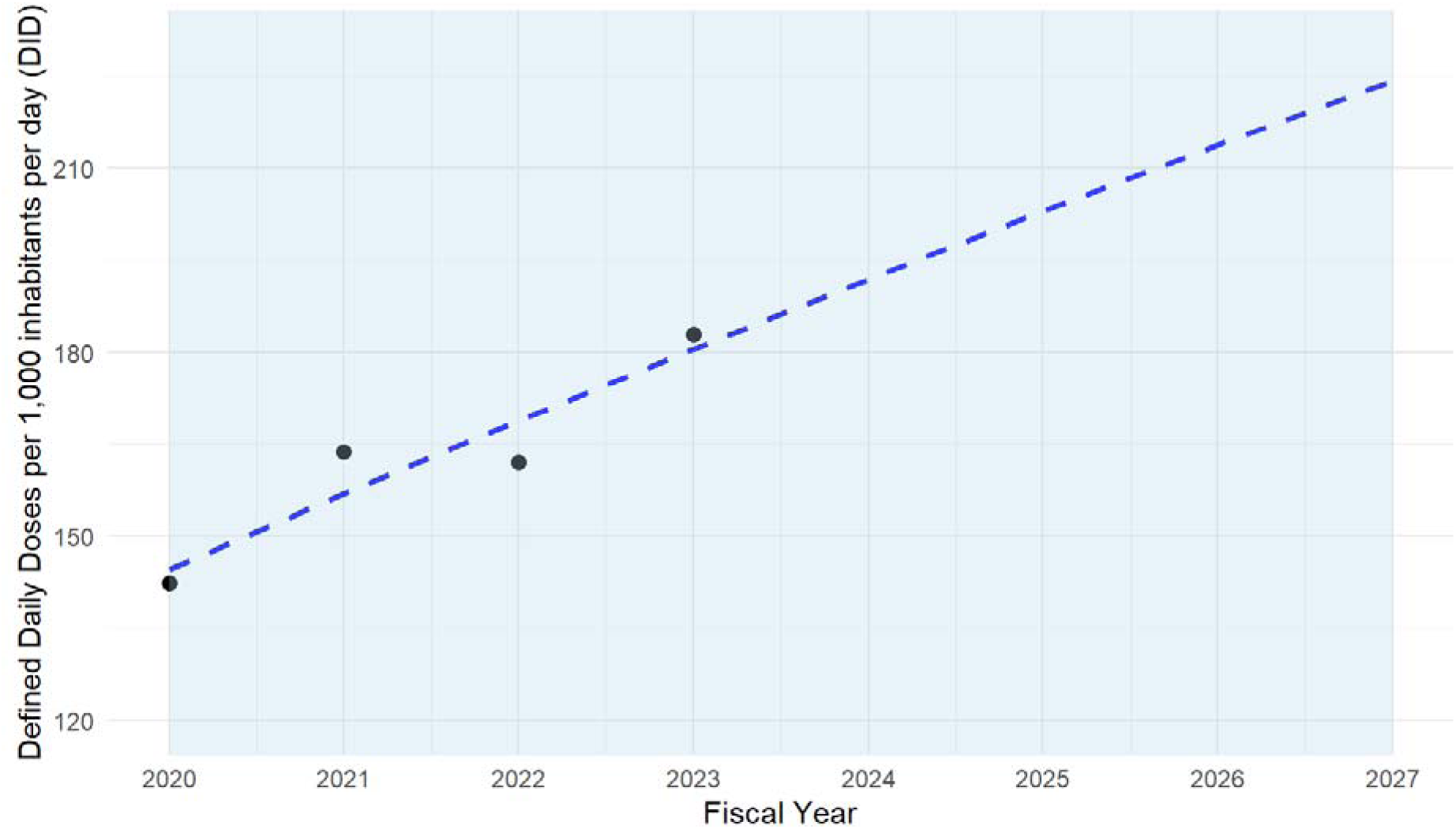
Forecast of national antibiotic consumption, Tanzania, 2020–2027. Observed and projected national antibiotic consumption (in DID) from 2020 to 2021 to 2026– 2027. The solid line represents the observed values (2020–2024), whereas the dashed line and shaded band show polynomial forecasted values with 95% confidence intervals. The projection indicates a continuing upward trend through 2027, reaching approximately 215 DID.

## Discussion

This study presents the first comprehensive, facility-level geospatial analysis of antibiotic consumption in Tanzania, utilizing eLMIS data from 2020 to 2024. The findings revealed marked regional and facility-level disparities, underscoring the need for context-specific AMS interventions. Similar spatial analyses have demonstrated that antibiotic use patterns vary substantially by geography and governance quality, influencing both access and resistance trends [4–6,13].

Antibiotic consumption increased nationally during the study period, with total DID rising from 142.4 in 2020–2021 to 182.8 in 2023–2024. This upward trend, which is consistent with our previous national analysis[3], reflects the persistently high demand for antibiotics across the healthcare system. Our analysis extends previous findings by mapping regional variations and identifying areas of concentrated use. High-consumption regions, such as Dar es Salaam, Ruvuma, and Lindi, contrast sharply with low-use areas, such as Katavi and Geita, suggesting heterogeneity in access, prescribing practices, and disease burden. This reinforces the need for AMS interventions tailored to regional consumption patterns rather than relying solely on national averages [1,4,7].

Dispensaries accounted for >70% of the total antibiotic distribution, underscoring the pivotal role of primary healthcare facilities in driving antibiotic use. This pattern confirms that lower-level facilities largely shape national antibiotic consumption. The predominance of dispensaries and health centers as antibiotic recipients reflects the accessibility and outpatient-driven nature of antimicrobial use in Tanzania. Similar findings were reported in multi-country analyses of primary care antibiotic use in sub-Saharan Africa, where lower-level facilities accounted for most outpatient antibiotic exposure [9,14]. Therefore, strengthening AMS at this level is essential, as most antibiotics are prescribed and dispensed outside the hospital setting. The relatively modest contributions of regional, zonal, and specialized hospitals likely reflect their smaller patient populations and tighter prescribing oversight [15,16].

When triangulated with molecule- and class-level findings, the results show that first-line agents, particularly penicillins, sulfonamide-trimethoprim combinations, macrolides, and fluoroquinolones, dominate across all tiers of care, mirroring national supply and importation patterns [8]. At the molecule level (ATC Level 5), amoxicillin and co-trimoxazole remain the leading agents, consistent with earlier longitudinal analyses of antibiotic utilization in Tanzania [3,9]. At higher classification levels (ATC Levels 3–4), the sustained dominance of broad-spectrum Access agents—penicillins with extended spectrum (J01CA), sulfonamide-trimethoprim combinations (J01EE), macrolides (J01FA), tetracyclines (J01AA), and fluoroquinolones (J01MA)— indicates continued reliance on a narrow set of therapeutic classes. This alignment across facility tiers and ATC levels highlights the structural determinants of antibiotic selection embedded within Tanzania’s Essential Medicines Framework. Integrating these insights into national AMS programs can refine procurement priorities, guide local formulary management, and focus surveillance on frequently used classes with the highest potential for resistance amplification. The inclusion of antibiotics, such as ciprofloxacin and azithromycin, among the most commonly used agents underscores the need to reinforce regulatory oversight and prescriber education [1].

The AWaRe analysis revealed high proportions of Access category antibiotics (≥60%) from 2021 onward, meeting the WHO global benchmark [1]. However, the persistent 35–40% Watch-antibiotic share highlights the continuing risk of resistance amplification and calls for sustained AMS efforts and surveillance. The negligible use of Reserve category antibiotics (<0.1%) is reassuring but may also indicate limited availability of critical second-line agents in some facilities, potentially constraining optimal care for severe infections [17]. Regions with low Access-category representation may likewise require targeted support to enhance equitable antibiotic availability, aligning AMS with the broader One Health and Universal Health Coverage goals [11].

Forecasting based on ARIMA and polynomial models suggests a continued upward trajectory in national antibiotic consumption through 2027, with an estimated 215 DID if the current patterns persist. This projected growth indicates that existing AMS interventions are insufficient to curb excessive antibiotic use. Embedding predictive analytics within Tanzania’s AMS framework could enable the earlier detection of consumption surges, support more accurate procurement planning, and guide proactive responses to emerging resistance hotspots [3].

Taken together, these findings highlight the value of integrating geospatial and data-driven surveillance into Tanzania’s AMR response framework. Mapping antibiotic consumption at the regional and facility levels enables evidence-based prioritization of interventions, particularly in the coastal and southern regions where use remains concentrated. Leveraging information from eLMIS for continuous monitoring can enhance resource allocation, improve procurement forecasting, and strengthen the design of AMS programs [3,7,15]. This approach aligns with WHO recommendations and offers a scalable model for other low- and middle-income countries seeking to implement context-specific AMS interventions [6].

The observed heterogeneity supports region-specific AMS actions under Tanzania’s 2020 National Action Plan, particularly through the integration of eLMIS-based consumption dashboards with regional health management teams. Future studies should extend this framework to include private sector facilities and link eLMIS data with microbiological resistance patterns to provide a more comprehensive view of antibiotic use and resistance dynamics.

### Limitations

This study has several limitations. First, the analysis relied on eLMIS facility-level supply data, which captured aggregated antibiotic distributions rather than patient-level prescriptions or actual use. Second, over 70% of the data originated from primary healthcare facilities, while regional, tertiary, and private hospitals were underrepresented, limiting the generalizability of the findings to higher-level and private care settings. Third, reporting from private and faith-based facilities within eLMIS remains incomplete, likely resulting in an underestimation of the overall national antibiotic consumption.

Despite these limitations, this dataset represents the most comprehensive and nationally representative evidence currently available on antibiotic supply and consumption trends in Tanzania, thus providing a strong basis for guiding AMS and supply chain policies.

## Conclusion

This national geospatial analysis revealed pronounced regional disparities in antibiotic consumption across Tanzania. Facility-level information from the Electronic Logistics Management Information System (eLMIS) revealed sustained growth in antibiotic use from 2020 to 2024, primarily driven by primary-care facilities and characterized by a small number of Access category antibiotics, notably amoxicillin and sulfamethoxazole-trimethoprim. The continued use of Watch agents highlights persistent gaps in prescribing oversight and the implementation of stewardship.

By combining spatial analytics with forecasting, this study demonstrates how digital health systems can generate actionable evidence for targeted antimicrobial stewardship interventions. These findings provide practical insights for strengthening Tanzania’s AMR Action Plan and present a scalable framework for other low- and middle-income countries seeking to balance equitable access, rational use, and resistance containment.

## Author Contributions

William Reuben: Conceptualization, Data collection, data validation, manuscript drafting Florah Boniface Makenya: data analysis, Validation data interpretation, manuscript editing,

Raphael Z. Sangeda: Conceptualization, data analysis, data interpretation, supervision, manuscript editing, corresponding author responsibilities

Daudi Ignasy Msasi: Data interpretation, supervision, manuscript editing.

Selesitine Ngoma: Data analysis, spatial mapping, supervision, manuscript review.

## Data Availability

The data supporting the findings of this study were sourced from the Tanzanian Ministry of Health and are accessible through the electronic Logistics Management Information System (eLMIS). These data are subject to restrictions owing to licensing and confidentiality agreements. Data access can be arranged through the authors, pending approval from the Ministry of Health.

## Acknowledgments

The authors thank the Ministry of Health, Tanzania, for access to the eLMIS data.

## Supplementary Tables

**Supplementary Table 1:** Regional antibiotic distribution intensity (DDD per 1,000 inhabitants per day), Tanzania, 2020–2024.

| Region | 2020–<br>2021 | 2021–<br>2022 | 2022–<br>2023 | 2023–<br>2024 | Region<br>Total | % of<br>Cumulative<br>Total | % Change<br>(2020–2021<br>to 2023–<br>2024) | Cumulative<br>% |
| --- | --- | --- | --- | --- | --- | --- | --- | --- |
| Dar es<br>Salaam | 15.37 | 8.25 | 4.98 | 9.36 | 37.95 | 5.8 | -39.1 | 5.8 |
| Ruvuma | 7.78 | 7.44 | 9.25 | 10.01 | 34.47 | 5.3 | 28.7 | 11.1 |
| Lindi | 4.38 | 11.00 | 9.58 | 9.04 | 34.00 | 5.2 | 106.7 | 16.3 |
| Iringa | 8.04 | 7.60 | 8.21 | 6.76 | 30.62 | 4.7 | -15.9 | 21.0 |
| Njombe | 7.60 | 8.13 | 6.68 | 7.33 | 29.74 | 4.6 | -3.5 | 25.6 |
| Mtwara | 4.71 | 9.39 | 7.37 | 7.75 | 29.22 | 4.5 | 64.7 | 30.1 |
| Dodoma | 6.70 | 7.01 | 7.80 | 7.51 | 29.02 | 4.5 | 12.1 | 34.6 |
| Tanga | 5.69 | 8.56 | 6.29 | 8.30 | 28.84 | 4.4 | 45.8 | 39.0 |
| Pwani | 7.64 | 7.18 | 6.69 | 7.26 | 28.77 | 4.4 | -4.9 | 43.4 |
| Kilimanjaro | 6.85 | 6.42 | 6.03 | 7.89 | 27.18 | 4.2 | 15.2 | 47.6 |
| Rukwa | 3.80 | 7.29 | 7.40 | 7.76 | 26.25 | 4.0 | 104.0 | 51.6 |
| Songwe | 5.65 | 6.24 | 7.46 | 6.56 | 25.92 | 4.0 | 16.1 | 55.6 |
| Kagera | 5.07 | 5.28 | 6.79 | 7.92 | 25.06 | 3.8 | 56.2 | 59.4 |
| Morogoro | 6.37 | 6.46 | 5.43 | 6.10 | 24.36 | 3.7 | -4.2 | 63.1 |
| Mbeya | 5.52 | 5.99 | 6.15 | 6.64 | 24.30 | 3.7 | 20.3 | 66.8 |
| Tabora | 3.95 | 5.73 | 6.28 | 6.69 | 22.65 | 3.5 | 69.3 | 70.3 |
| Mara | 3.69 | 4.64 | 6.64 | 7.20 | 22.17 | 3.4 | 95.4 | 73.7 |
| Manyara | 3.99 | 5.82 | 5.62 | 6.63 | 22.05 | 3.4 | 66.2 | 77.1 |
| Singida | 4.67 | 5.56 | 4.88 | 6.80 | 21.91 | 3.4 | 45.6 | 80.5 |
| Mwanza | 5.04 | 4.80 | 5.44 | 6.57 | 21.85 | 3.4 | 30.3 | 83.9 |
| Arusha | 3.97 | 5.64 | 4.85 | 6.35 | 20.81 | 3.2 | 60.2 | 87.1 |
| Kigoma | 4.31 | 4.65 | 5.23 | 6.37 | 20.57 | 3.2 | 47.6 | 90.3 |
| Shinyanga | 3.34 | 4.57 | 5.01 | 5.53 | 18.44 | 2.8 | 65.4 | 93.1 |
| Simiyu | 2.71 | 3.85 | 4.15 | 4.95 | 15.66 | 2.4 | 82.9 | 95.5 |
| Geita | 2.82 | 3.46 | 3.86 | 5.30 | 15.44 | 2.4 | 87.7 | 97.9 |
| Katavi | 2.73 | 2.81 | 3.93 | 4.21 | 13.68 | 2.1 | 54.4 | 100.0 |
| **National**<br>**Total** | **142.37** | **163.76** | **162.01** | **182.79** | **650.92** | **100.0** | **--** | **--** |

*Note: Values show regional sums of defined daily doses (DDD) per 1,000 inhabitants per day (DID) based on eLMIS antibiotic supply data aggregated by fiscal year. The region total represents the cumulative DID across 2020–2024 and the percentage change reflects the relative change between the first and last fiscal years.*

**Supplementary Table 2:** Top antibiotics by consumption (DDD per 1,000 inhabitants per day), Tanzania, 2020–2024.

| Antibiotic (ATC code) | 2020–<br>2021 | 2021–<br>2022 | 2022–<br>2023 | 2023–<br>2024 | Cumulative<br>Total | % of Cumulative<br>Total |
| --- | --- | --- | --- | --- | --- | --- |
| Amoxicillin (J01CA04) | 41.53 | 39.62 | 29.69 | 34.08 | 144.91 | 22.3 |
| Sulfamethoxazole +<br>Trimethoprim (J01EE01) | 44.91 | 38.12 | 26.78 | 30.70 | 140.51 | 21.6 |
| Doxycycline (J01AA02) | 7.63 | 20.37 | 23.28 | 19.04 | 70.32 | 10.8 |
| Ciprofloxacin (J01MA02) | 12.56 | 17.58 | 20.16 | 19.78 | 70.08 | 10.8 |
| Erythromycin (J01FA01) | 10.86 | 14.97 | 15.07 | 14.28 | 55.17 | 8.5 |
| Metronidazole (J01XD01) | 6.48 | 10.38 | 16.66 | 20.48 | 53.99 | 8.3 |
| Ampicillin + Cloxacillin<br>(J01CA51) | 6.92 | 7.67 | 11.58 | 16.11 | 42.27 | 6.5 |
| Azithromycin (J01FA10) | 1.05 | 3.39 | 6.67 | 12.08 | 23.19 | 3.6 |
| Amoxicillin + Clavulanic<br>Acid (J01CR02) | 1.40 | 2.36 | 3.26 | 3.19 | 10.21 | 1.6 |
| Ceftriaxone (J01DD04) | 1.26 | 1.62 | 1.35 | 2.94 | 7.18 | 1.1 |
| Phenoxymethyl Penicillin<br>(J01CE02) | 1.91 | 2.04 | 2.13 | 0.30 | 6.38 | 1.0 |

| **Antibiotic (ATC code)** | **2020–**<br>**2021** | **2021–**<br>**2022** | **2022–**<br>**2023** | **2023–**<br>**2024** | **Cumulative**<br>**Total** | **% of Cumulative**<br>**Total** |
| --- | --- | --- | --- | --- | --- | --- |
| Cephalexin (J01DB01) | 0.84 | 0.67 | 1.23 | 2.21 | 4.93 | 0.8 |
| Benzyl Penicillin<br>(J01CE01) | 1.34 | 1.37 | 0.88 | 1.32 | 4.90 | 0.8 |
| Gentamicin (J01GB03) | 0.12 | 0.76 | 0.86 | 0.89 | 2.63 | 0.4 |
| Nitrofurantoin (J01XE01) | 0.20 | 0.42 | 0.54 | 1.33 | 2.49 | 0.4 |
| Ciprofloxacin +<br>Tinidazole (J01RA11) | 1.48 | 0.11 | 0.01 | 0.00 | 1.60 | 0.2 |
| Tinidazole (J01XD02) | 0.22 | 0.13 | 0.05 | 1.13 | 1.53 | 0.2 |
| Cefixime (J01DD08) | 0.04 | 0.05 | 0.45 | 0.85 | 1.40 | 0.2 |
| Ampicillin (J01CA01) | 0.03 | 0.35 | 0.41 | 0.56 | 1.35 | 0.2 |
| Flucloxacillin +<br>Amoxicillin (J01CF05) | 0.25 | 0.27 | 0.39 | 0.44 | 1.35 | 0.2 |
| **Subtotal (top 20)** | **144.64** | **173.09** | **175.99** | **187.52** | **681.24** | **≈ 99 % of total**<br>**consumption** |

Note:
The following antibiotics were recorded to have negligible or no distribution during the study period (2020–2024): kanamycin (J01GB04), clindamycin (J01FF01), nalidixic acid (J01MB02), ceftazidime (J01DD02), tazobactam (J01CG02), and amikacin (J01GB06).

**Supplementary Table 3:** ATC Level 4 antibiotic classes--annual totals of defined daily doses per 1,000 inhabitants per day (DID) by fiscal year (2020–2024) and cumulative class totals. Values derived from eLMIS facility-level supply data. The Class Total” rows represent the national sum per year; the rightmost column provides the cumulative DID per class across all years.

| **Level 4 Antibiotic (class)** | **Fiscal Year** |  |  |  | **Class**<br>**Total** |
| --- | --- | --- | --- | --- | --- |
|  | **2020-2021** | **2021-2022** | **2022-2023** | **2023-**<br>**2024** |  |
| Amphenicols (J01BA) | 0.009165 | 0.014371 | 0.013387 | 0.012936 | 0.049859 |
| Beta-lactamase inhibitors<br>(J01CG) |  |  | 0 | 0 | 0 |
| Beta-lactamase resistant<br>penicillins (J01CF) | 0.279362 | 0.269429 | 0.395662 | 0.440577 | 1.38503 |
| Beta-lactamase sensitive penicillins (J01CE) | 3.603444 | 3.581722 | 3.024911 | 1.618172 | 11.828249 |
| Carbapenems (J01DH) | 0.00058 | 0.00931 | 0.014997 | 0.015757 | 0.040644 |
| Combinations of antibacterials (J01RA) | 1.485495 | 0.123262 | 0.006455 | 0.002949 | 1.618161 |
| Combinations of penicillins, incl. beta-lactamase inhibitors (J01CR) | 1.403461 | 2.364825 | 3.255315 | 3.186898 | 10.210499 |
| Combinations of sulphonamides and trimethoprim incl. derivatives (J01EE) | 44.909796 | 38.118795 | 26.783921 | 30.695668 | 140.50818 |
| First generation cephalosporins (J01DB) | 0.838575 | 0.674016 | 1.227325 | 2.206148 | 4.946064 |
| Fluoroquinolones (J01MA) | 12.976689 | 17.957546 | 20.340317 | 19.861126 | 71.135678 |
| Fourth generation cephalosporins (J01DE) | 0.088864 | 0.043165 | 0.055327 | 0.212251 | 0.399607 |
| Glycopeptide antibacterials (J01XA) | 0.000233 | 0.000713 | 0.00096 | 0.006894 | 0.0088 |
| Imidazole derivatives (J01XD) | 6.887814 | 11.194347 | 16.704434 | 21.605792 | 56.392387 |
| Lincosamides (J01FF) | 0.002429 | 0.000777 | 0.00135 | 0.000667 | 0.005223 |
| Macrolides (J01FA) | 11.995891 | 18.38516 | 21.948916 | 26.925127 | 79.255094 |
| Nitrofurans derivatives (J01XE) | 0.198007 | 0.418473 | 0.535158 | 1.333722 | 2.48536 |
| Other aminoglycosides (J01GB) | 0.120192 | 0.758478 | 0.86349 | 0.887055 | 2.629215 |
| Other antibacterials (J01XX) | 0.07758 | 0.157961 | 0.000084 | 0.048866 | 0.284491 |
| Other quinolones (J01MB) | 0 | 0 | 0 | 0 | 0 |
| Penicillins with extended spectrum (J01CA) | 48.484802 | 47.63442 | 41.66827 | 50.750147 | 188.537639 |
| Second-generation cephalosporins (J01DC) | 0.008357 | 0 | 0.004774 | 0.018324 | 0.031455 |
| Streptomycins (J01GA) | 0.000051 |  | 0.000201 |  | 0.000252 |
| Tetracyclines (J01AA) | 7.63499 | 20.368453 | 23.27892 | 19.035823 | 70.318186 |
| Third-generation cephalosporins (J01DD) | 1.361968 | 1.681346 | 1.886132 | 3.925111 | 8.854557 |
| **Class Total** | **142.367745** | **163.756569** | **162.010306** | **182.79001** | **650.92463** |

**Supplementary Table 4:** Forecast summary of national antibiotic consumption (defined daily doses per 1,000 inhabitants per day, DID), Tanzania, 2020–2027 (ARIMA and polynomial projections).

| **Fiscal Year** | **Observed DID** | **ARIMA Forecast** | **Polynomial Forecast** | **95% Confidence Interval (ARIMA)** |
| --- | --- | --- | --- | --- |
| 2020–2021 | 142.37 | -- | -- | -- |
| 2021–2022 | 163.76 | -- | -- | -- |
| 2022–2023 | 162.01 | -- | -- | -- |
| 2023–2024 | 182.79 | -- | -- | -- |
| 2024–2025 | -- | 193.4 | 195.6 | 188.1 – 198.7 |
| 2025–2026 | -- | 202.5 | 205.8 | 194.3 – 210.6 |
| 2026–2027 | -- | 211.1 | 215.2 | 200.4 – 222.8 |

Note: Forecasts derived from national annual DID values (2020–2024) using ARIMA (1,1,0) and second-degree polynomial models. Both models predicted continued growth in antibiotic consumption through 2027, with the polynomial model providing a marginally better fit ( $R^2 = 0.97$ ).

